# Strength training rescues mitochondrial dysfunction in skeletal muscle of patients with myotonic dystrophy type 1

**DOI:** 10.1101/2023.01.20.23284552

**Authors:** Valeria Di Leo, Conor Lawless, Marie-Pier Roussel, Tiago B. Gomes, Gráinne S. Gorman, Oliver M. Russell, Helen A. L. Tuppen, Elise Duchesne, Amy E. Vincent

**Affiliations:** Wellcome Centre for Mitochondrial Research, Translational and Clinical Research Institute, Faculty of Medical Sciences, Newcastle University, UK; Department of Fundamental Sciences, Université du Québec à Chicoutimi, Québec, Canada; NHS Highly Specialised Service for Rare Mitochondrial Disorders, Royal Victoria Infirmary, Newcastle Upon Tyne, UK; Department of Health Sciences, Université du Québec à Chicoutimi, Québec, Canada

**Author notes:** Corresponding Authors: Prof Elise Duchesne, PT, PhD, Department of Health Sciences, Université du Québec à Chicoutimi, Chicoutimi, Québec, Canada, Email address, Dr. Amy E. Vincent, PhD, Wellcome Centre for Mitochondrial Research, 4^th^ Floor Cookson Building, Framlington Place, Newcastle University, Newcastle upon Tyne, Tyne and Wear, UK. These authors contributed equally to the manuscript (Elise Duchesne, Amy E. Vincent).

**Keywords:** myotonic dystrophy type 1, skeletal muscle, mitochondrial dysfunction, oxidative phosphorylation deficiency, strength training, myotonic dystrophy type 1 therapy

## Abstract

Myotonic dystrophy type 1 (DM1) is a neuromuscular disorder, for which no cure exists. This study investigates the effects of 12-week strength training on mitochondrial oxidative phosphorylation in skeletal muscle in a cohort of DM1 patients (n=11, males) in comparison to untrained sex-matched healthy subjects. Immunofluorescence was used to assess protein levels of key respiratory chain subunits of complex I (CI) and complex IV (CIV), and markers of mitochondrial mass and cell membrane in individual myofibers sampled from biopsies. We classified each patient myofiber as having normal, low or high levels of CI and CIV and compared the proportions of affected fibers before and after exercise training. The significance of changes observed between pre- and post-exercise training within patients was estimated using a permutation test.

At baseline, DM1 patients present with significantly decreased mitochondrial mass, and isolated or combined CI and CIV deficiency. After strength training, in most patients a significant increase in mitochondrial mass was observed, and all patients showed a significant increase in CI and/or CIV protein levels. Remarkably, 12-week strength training is sufficient to partially rescue mitochondrial dysfunction in DM1 patients, suggesting exercise as an inexpensive and accessible therapy option.

## 1. Introduction

Myotonic dystrophy type 1 (DM1) is a dominant autosomal disorder that affects 1 in 20,000 worldwide [1]. Population incidences vary in different parts of the world, reaching a higher frequency of 1 in 475 in the Québec region of Saguenay-Lac-Saint-Jean (Canada) [2]. DM1 etiology is explained by the toxic gain of function of the Dystrophia Myotonica Protein Kinase (DMPK), which originates from a CTG triplet repeat expansion in the 3’ untranslated region of the *DMPK* gene [3, 4]. The CTG triplet repeat microsatellite region contains between 5 and 37 repeats in non-DM1 individuals [5, 6]. In DM1 patients however, the expansion may range from 50 to thousands of repeats and further expand in post-mitotic cells, such as skeletal muscle (SKM), presenting as somatic mosaicism [7, 8]. Depending on the number of CTG triplet repeats inherited, the age of onset can vary between congenital, childhood, juvenile, adult, or late [9]. Although presenting as a systemic disease, the most prominent symptoms associated with DM1 affect the SKM apparatus with weakness, myotonia and atrophy [10].

The DMPK gain of function triggers RNA toxicity, characterized by nuclear sequestration of transcription and splicing factors that eventually dysregulate the downstream alternative splicing machinery [11]. Traditionally, research has focused on understanding RNA toxicity and reversing the mechanisms of the aberrant splicing program. However, a myriad of proteins and mechanisms are affected in DM1 pathology, including nuclear and cytosolic alterations, but also organelle-specific perturbations, such as mitochondria [12]. DMPK has been found to specifically bind to the outer mitochondrial membrane [13], and its overexpression in myoblasts induces fragmentation and perinuclear clustering of mitochondria, resulting in the increase of both autophagy and apoptosis [14]. Moreover, DMPK interacts with tyrosine kinase Src and Hexokinase-II in the formation of a multimeric complex on the outer mitochondrial membrane, which orchestrates a fine-tuned regulation of intracellular oxidative stress and pro-survival processes via glucose starvation stimuli [15-17]. Strikingly, there is evidence that DM1 patients present with different mitochondrial morphology, dynamics and function affecting the structure of the SKM sarcoplasmic reticulum with subsequent mitochondrial aggregation [18], metabolic impairment [19] and neuromuscular junction alterations [20, 21]. More recent *in vivo* studies demonstrated that DM1 patients present with oxidative metabolism impairment in both SKM and brain [22]. Like mitochondrial myopathy patients, DM1 patients present with elevated FGF21 serum levels due to insulin resistance and mitochondrial dysfunction [23, 24]. Furthermore, it was recently demonstrated that metformin treatment could improve mobility in DM1 patients; specifically, in DM1-derived fibroblasts, it reverses the impaired metabolism and mitochondrial dysfunction [25, 26].

Although DM1 is one of the most common adult-onset neuromuscular disorders, there is no cure or treatment available. Many different approaches, including the genomic correction of the *DMPK* gene, the generation of antisense oligonucleotides for the alternative splicing correction and the use of small molecules for the modulation of signaling pathways, have been extensively explored to treat DM1 pathology and different clinical trials are underway [27, 28]. At present, the best option for DM1 patients is to provide symptomatic therapy when needed, to improve quality of life and increase life expectancy. To this end, strength training has been tested as an accessible, low-cost way to improve SKM weakness and increase muscle strength by inducing hypertrophy in DM1 patients [29-35]. Particularly in mitochondrial myopathy patients, 12-week strength training was demonstrated to induce amelioration in the oxidative phosphorylation (OXPHOS) defects, which mainly cause myopathy and weakness [36]. Mitochondrial OXPHOS deficiency has been demonstrated in DM1 patients compared to healthy individuals [37]. However, the effects of strength training on mitochondrial OXPHOS deficiency have never been assessed before in SKM tissue from DM1 patients. The aim of this study is to investigate mitochondrial dysfunction and whether it improved in a cohort of DM1 patients, who underwent a 12-week strength training program [34]. We investigate whether DM1 patients present with OXPHOS defects at a baseline level in SKM tissue, and whether 12-week strength training induces any changes in mitochondrial function.

## 2. Materials and Methods

### 2.1 Experimental design

This study is a secondary analysis of a larger project, in which DM1 patients (n=11) were recruited to participate in a 12-week strength training program [34]. DM1 patients were between 31 and 60 years old, were able to walk independently and to give their informed consent. Vastus lateralis muscle biopsies from healthy controls (n=3) and DM1 patients pre- and post-exercise (**Table 1**) were collected at the Université du Québec à Chicoutimi (Canada), as previously reported [34]. Biopsy sections (10µm) were cut on glass slides at the Université du Québec (Chicoutimi, Canada). Samples were stored on dry ice until shipment to Newcastle University (Newcastle upon Tyne, United Kingdom). Upon receipt, samples were stored at - 80°C.

**Table 1:**
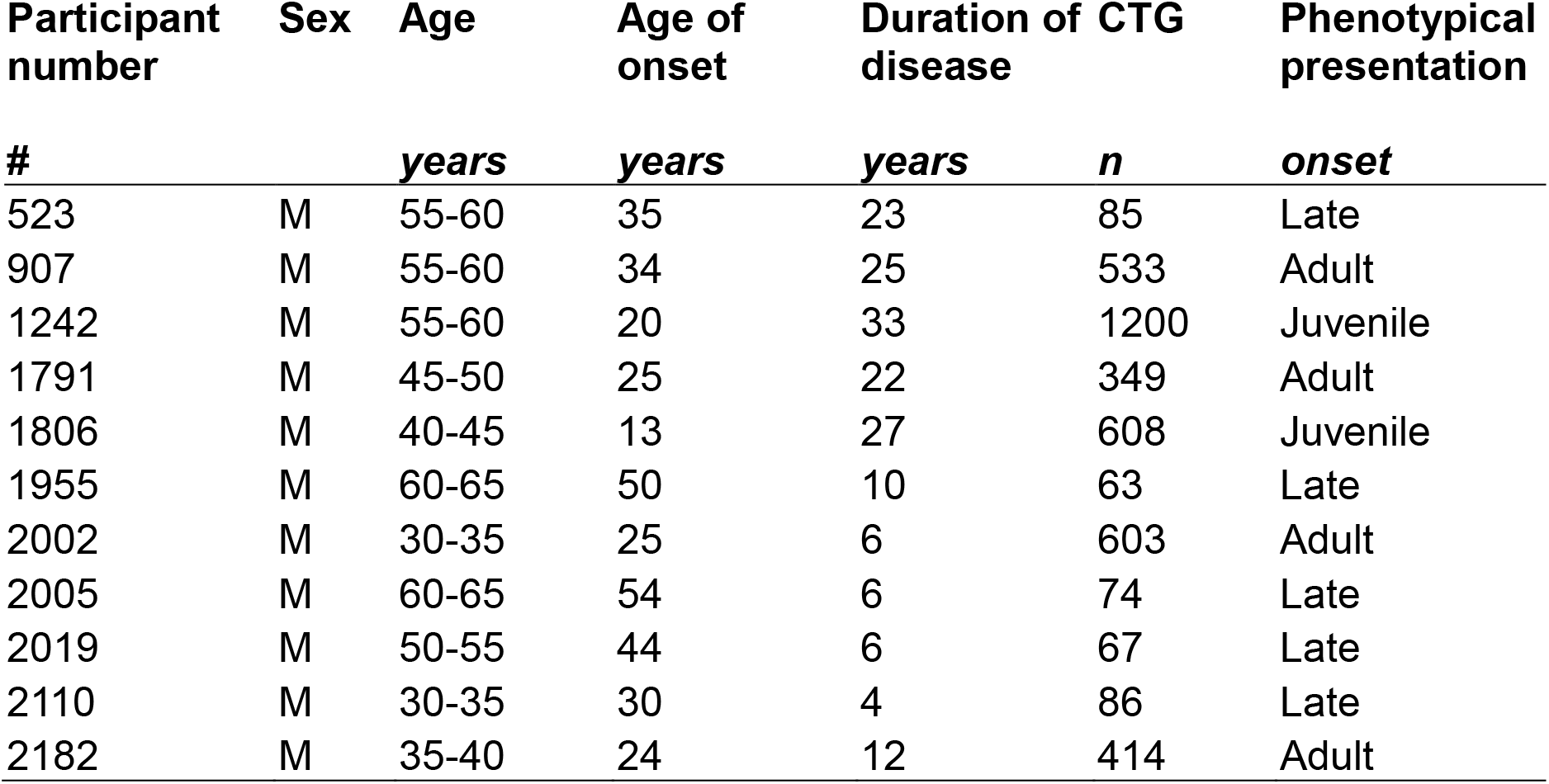
Characteristics of participants enrolled in the strength training program. The following details are listed: participant number, sex, age, age of onset of the disease, duration of disease since diagnosis or first symptoms appearance, number of CTG triplet repeats expansion measured in blood, and phenotypical presentation.

### 2.2 Clinical measurements

All clinical measurements related to the DM1 cohort were previously published [34]. The number of CTG triplet repeats in blood were retrieved from medical files. Age refers to the one at the time of recruitment for strength training program. Age of onset refers to when first symptoms appeared and/or when a genetic test was performed for diagnosis purposes. Duration of disease was calculated as the difference between age of patients and age of onset of the disease. Phenotypical presentation was classified based on the number of CTG triplet repeats and age of onset.

### 2.3 Quadruple immunofluorescence

SKM sections were stained for quadruple immunofluorescence (QIF) to assess OXPHOS [38]. The staining included the immunolabelling of key subunit NADH:Ubiquinone Oxidoreductase Subunit B8 (NDUFB8) for CI and Mitochondrially Encoded Cytochrome C Oxidase I (COX1) for CIV, together with Voltage Dependent Anion Channel 1 (VDAC1) as a mitochondrial mass marker and Laminin subunit alpha-1 (LAMA1) as a cell membrane marker.

### 2.4 Fluorescent microscopy

Fluorescent images were taken with Zeiss Cell Discoverer 7 (CD7) and analyzed using Zen 2011 (black edition) software. The CD7 includes the following parts: a Hamamatsu Fusion and a Zeiss Axiocam 506 monochrome camera; 5x/0.35, 20x/0.7 and 50x/1.2NA lenses; a Zeiss LED light source (Colibri 7). Image acquisition was performed at 20x magnification using a motorized stage AxioImager M1 and the tiling function in Zen software. For each section, a .czi file was generated using the stitching function in Zen.

### 2.5 Image analysis

Stitched images of the SKM sections (.czi files) were analyzed using *Quadruple Immuno Analyser*, an in-house software written in MatLab R2015a [38]. The software automatically created a segmentation map of the SKM fibers’ boundaries using LAMA1 signal. The mean signal intensities for each channel in each single SKM fiber were exported in tabular format as .csv files.

### 2.6 Linear regression and 95% predictive interval model

Statistical analysis of QIF data was conducted using a 95% predictive interval linear regression model based on combined control population of fibers, as previously described [39]. In patient samples, all fibers lying within the control predictive interval were classified as normal for NDUFB8 or COX1 (fibers^normal^). Fibers below the interval were classified as fibers with a low level (fibers^low^), and above the interval as fibers with a high level (fibers^high^). The relationships between VDAC1 and NDUFB8 or COX1, respectively, for each patient and all controls are shown as scatterplots, here referred as 2DMito plots (https://github.com/VDLNCL/DM1-mitochondria/blob/main/Report.pdf).

### 2.7 Bootstrapping and permutation test

We used statistical bootstrapping to estimate uncertainty about the proportions of fibers classified as fibers^normal^, fibers^low^ and fibers^high^. We used the permutation test to estimate whether observed differences in proportions after strength training are significant. The number of resamples was set as N=5,000. We also generated bootstrapping estimates of Δfibers^low^ and Δfibers^high^, the differences in proportion of fibers in each class (post-exercise - pre-exercise) to quantify our uncertainty about changes after exercise. A permutation test was performed for each class of fibers in each patient to calculate whether exercise-induced changes were statistically significant using the null hypothesis that the labels “pre-exercise” and “post-exercise” are interchangeable. The permutation test provided p-values through a one-sided t-test. Δfibers^low^ for both NDUFB8 and COX1 was classified as significant when there was a decrease after exercise (Δfibers^low^ < 0) and p<0.05. Δfibers^high^ for both NDUFB8 and COX1 was classified as significant when there was an increase after exercise (Δfibers^high^ > 0) and the p-value was <0.05. All p-values were corrected for multiple testing by controlling the False Discovery Rate.

### 2.8 Statistical tests

All statistical tests were performed in R (version 3.5.2, https://intro2r.com/citing-r.html). A t-test was used to assess the difference between the population means for VDAC1. The function *t*.*test* was used to perform two-sided t-test. An ANOVA test was used to test any significant difference in the mean of independent populations (control and pre-exercise populations, or pre- and post-exercise populations). The function *aov* was used to perform two-way ANOVA test. An over-representation test was used to test the statistical over-representation of a subset of observations (COX1^low^ or COX1^high^) within a subset of collected data (NDUFB8^low^ or NDUFB8^high^). The function *hypertest* (https://github.com/VDLNCL/DM1-mitochondria/blob/main/Over-representation%20test.R) was used to perform the over-representation test, where *x* was defined as number of NDUFB8^low^&COX1^low^ or NDUFB8^high^&COX1^high^, *m* as number of COX1^low^ or COX1^high^, *y* as number of NDUFB8^low^ or NDUFB8^high^, *n* as the total number of fibers per sample.

## 3. Results

### 3.1 DM1 cohort genotypic and phenotypical presentations

The cohort of subjects used in this study included DM1 participants (n=11, male) (**Table 1**) and healthy sex-matched controls (n=3, male, age 33.7±9 years). The DM1 cohort presents with a mean age of 48±10.6 years, and a mean age of onset of 32.2±12.2 years. The CTG triplet repeat expansion varies with a minimum in patient #1955 (n=63) and a maximum in patient #1242 (n=1,200). As expected, the CTG triplet repeat expansion shows a strong negative correlation with the age of onset displaying a slope significantly different from zero (Pearson, R= -0.7; p-value = 0.016, **Figure 1**). The phenotypical presentations observed in the DM1 cohort were juvenile (n=2), adult (n=4) and late (n=5) onset (**Table 1**), according to previous publications [9, 34]. Importantly, duration of disease varies extremely across the DM1 cohort with more than 20 years in some patients (min=22, max=33, n=5) or less than 20 years in others (min=4, max=12, n=6) (**Table 1**).

**Figure 1:**
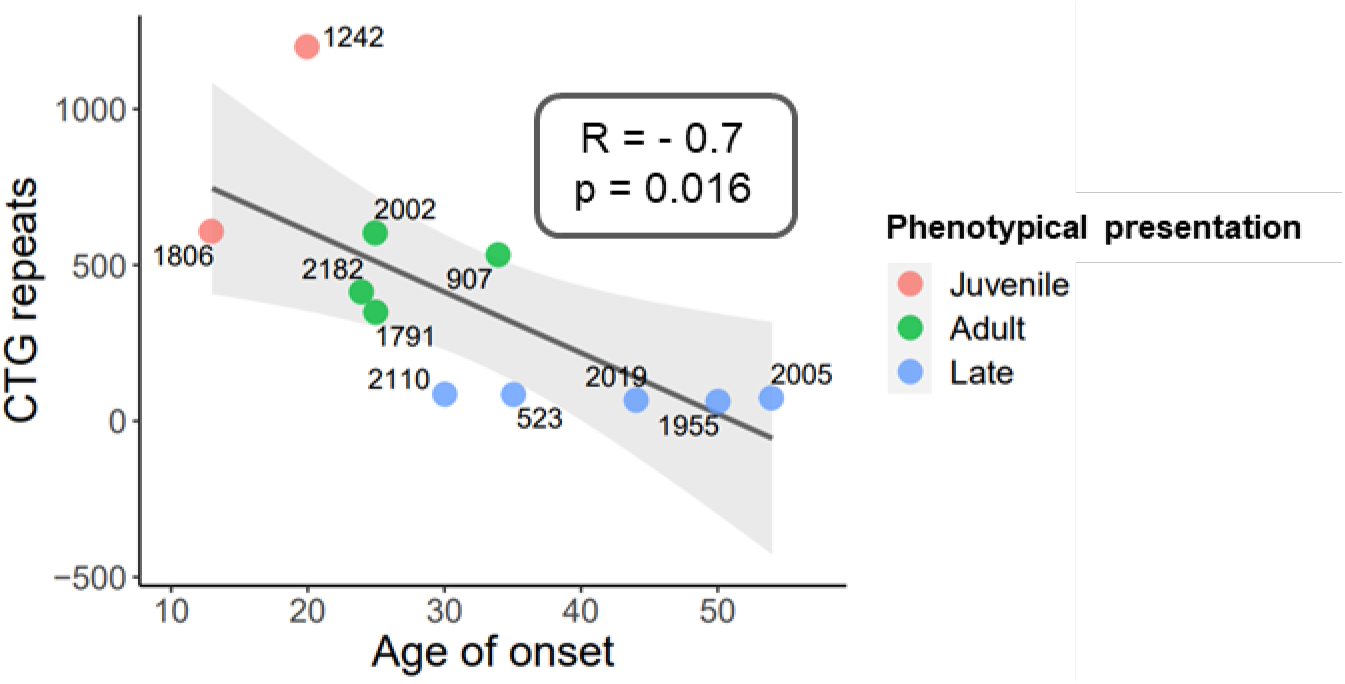
Genotypic and phenotypical presentation of DM1 cohort. The CTG triplet repeat expansion negatively correlates with the age of onset of DM1 disease, resulting in three phenotypical presentation classes: juvenile, adult, and late onset. Correlation coefficient (R) and p-value are indicated.

### 3.2 Mitochondrial mass deficiency and changes after strength

QIF labelling of NDUFB8, COX1, VDAC1 and LAMA1, was used to investigate mitochondrial mass and OXPHOS defects in the SKM biopsies of the DM1 cohort. QIF staining was not possible in pre-exercise section of case #2002, so the patient was excluded from further analyses. A representative image of the QIF staining is provided in **Figure 2**. VDAC1 intensities from single myofibers in each section were assessed to calculate a mean value for each pre- and post-exercise sample across the DM1 cohort [39]. The DM1 pre-exercise group (mean=5979.1) shows significantly decreased mitochondrial mass compared to healthy controls (mean=6591.6) (two-way ANOVA test, p-value < 0.0001, **Figure 3A**). No difference is observed between the pre-exercise group and post-exercise group (mean=5998.7) (two-way ANOVA test, **Figure 3A**). When comparing each pair of pre- and post-exercise samples, six of 10 patients exhibit a significant increase in VDAC1 mean value (t-test two-sided, p-value < 0.05, **Figure 3B**). However, four patients (cases #2182, #1955, #2005 and # 2019) present with a significant decrease in mitochondrial mass after exercise (t-test two-sided, p-value < 0.05, **Figure 3B**).

**Figure 2:**
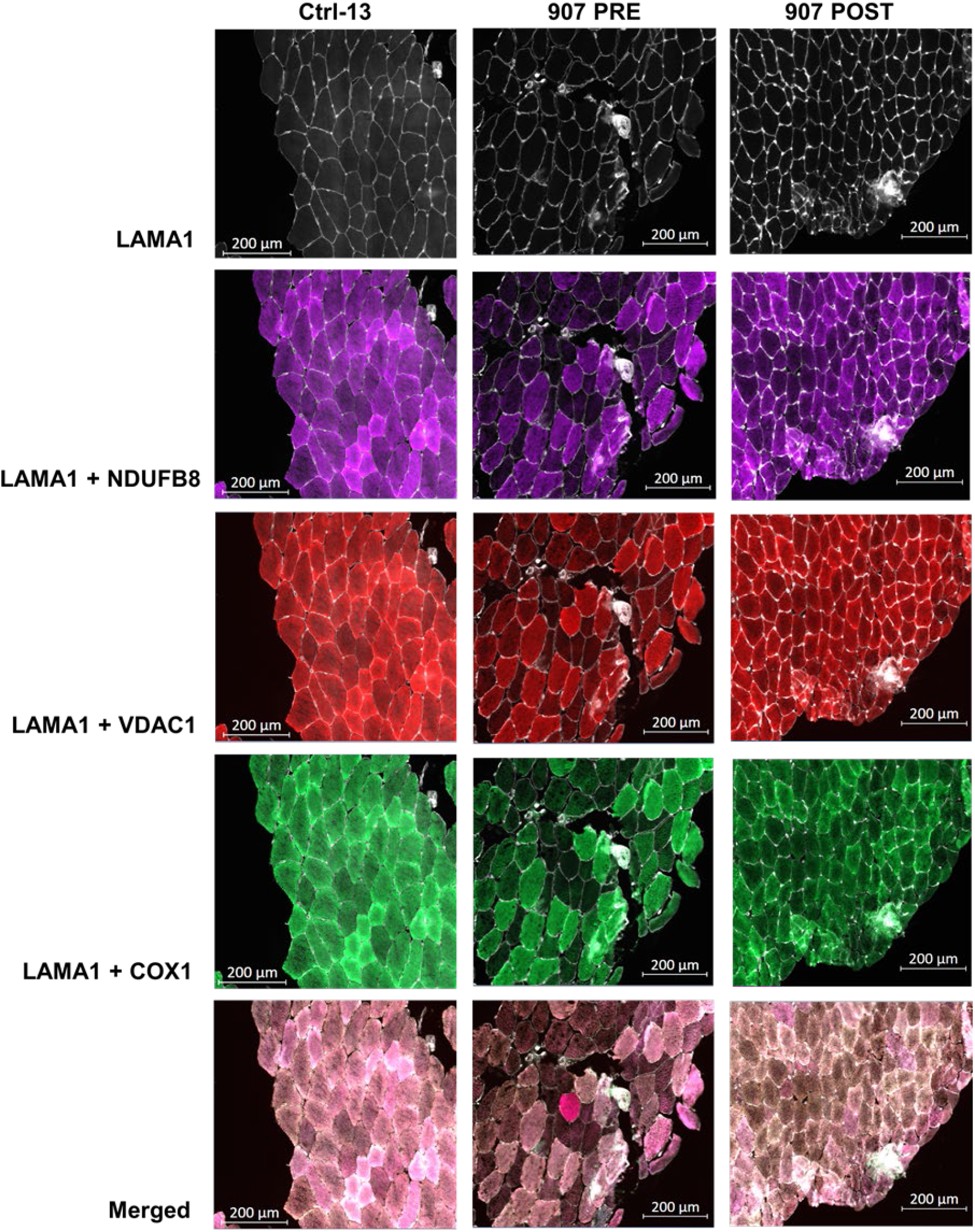
Representative QIF images of control subject, pre- and post-exercise pair from DM1 cohort after strength training. Representative images of quadruple immunofluorescence staining for LAMA1 (membrane marker), VDAC1 (mitochondrial mass marker), NDUFB8 (complex I marker) and COX1 (complex IV marker) in SKM sections from healthy Ctrl-13, P907 PRE (pre-exercise) and P907 POST (post-exercise) biopsies. The last row is the merge of all channels.

**Figure 3:**
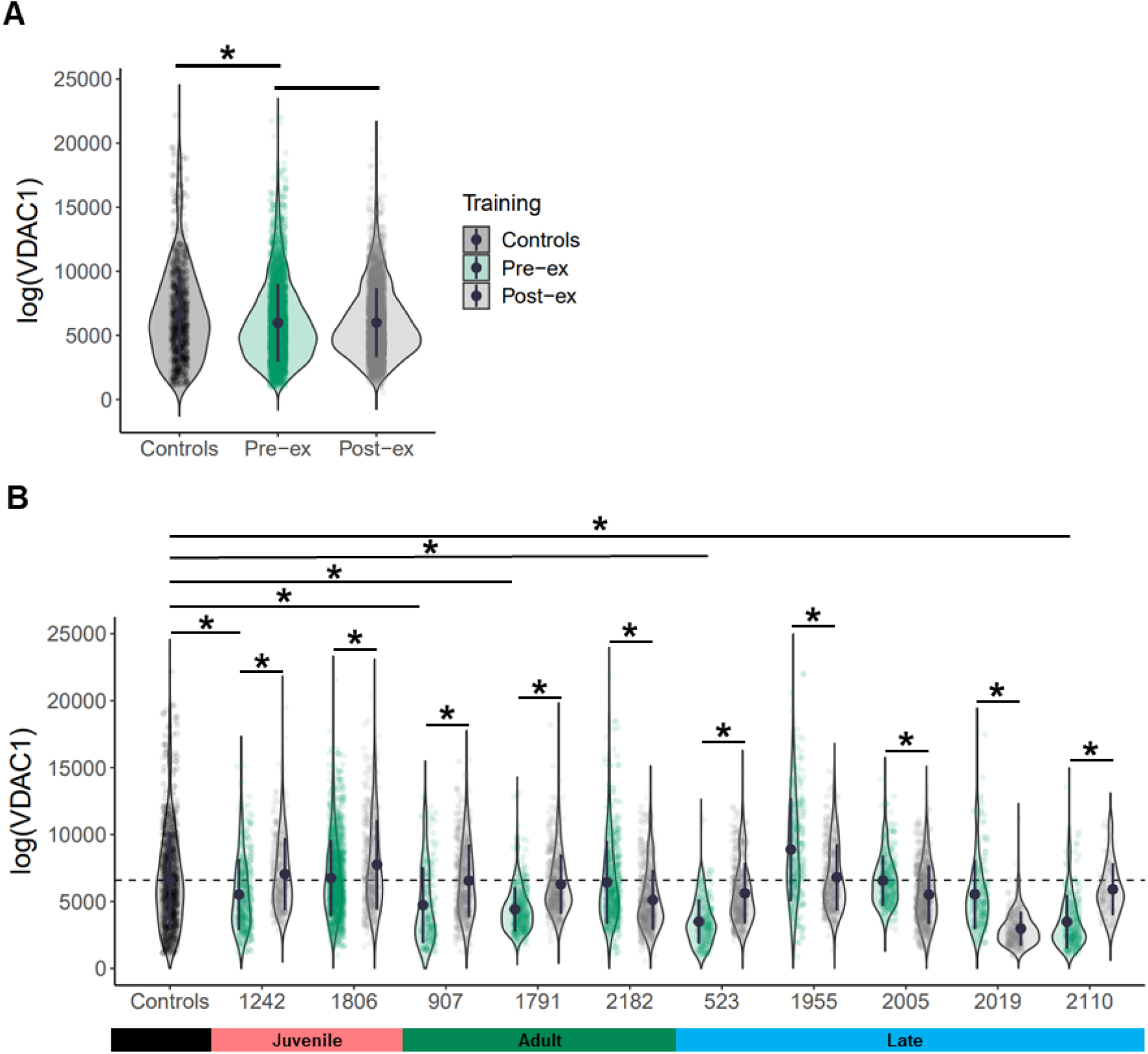
Mitochondrial mass and exercise-induced effects in DM1 cohort. VDAC1 values were compared between pre-exercise group and controls or post-exercise group, respectively. Mean values, two-way ANOVA test, p-value < 0.05 (*) (A). Mean value of mitochondrial mass of controls fibers is indicated as dashed black line. VDAC1 mean values were compared between each pre-exercise case and grouped controls. VDAC1 mean values were compared between pre- and post-exercise samples for each patient. Whiskers indicate the min and max values. Mean values, t-test two-sided, p-value < 0.05 (*) (B).

### 3.3 CI and CIV deficiency in DM1 patients

We classified myofibers after QIF staining using a linear regression between VDAC1 intensity and either NDUFB8 or COX1, using data from control subjects. Patient’ fibers were classified by comparison with the 95% predictive interval from the regression. A typical output of the statistical analysis is shown in **Supplementary Figure 1**. Percent of fibers below and above the control population, NDUFB8^low^ and COX1^low^, NDUFB8^high^ and COX1^high^ fibers respectively, were calculated for each pair of samples for each patient in the DM1 cohort (**Table 2**). At a baseline level, only patient #523 presents with 0% NDUFB8^low^ fibers. The rest of the DM1 cohort present with NDUFB8^low^ fibers, although patients #1242, #1791, #2182 and #2005 display less than 5% fibers classified as NDUFB8^low^ (**Figure 4A**). Similarly, five of 10 patients show a high proportion of COX1^low^ fibers (#1242, #907, #523, #1955 and #2019, (**Figure 4C**). The proportion of NDUFB8^high^ fibers is high in patient #523 before exercise (**Figure 4B**), whereas the proportion of COX1^high^ fibers is close to 0% in most patients before exercise (**Figure 4D**).

**Table 2:**
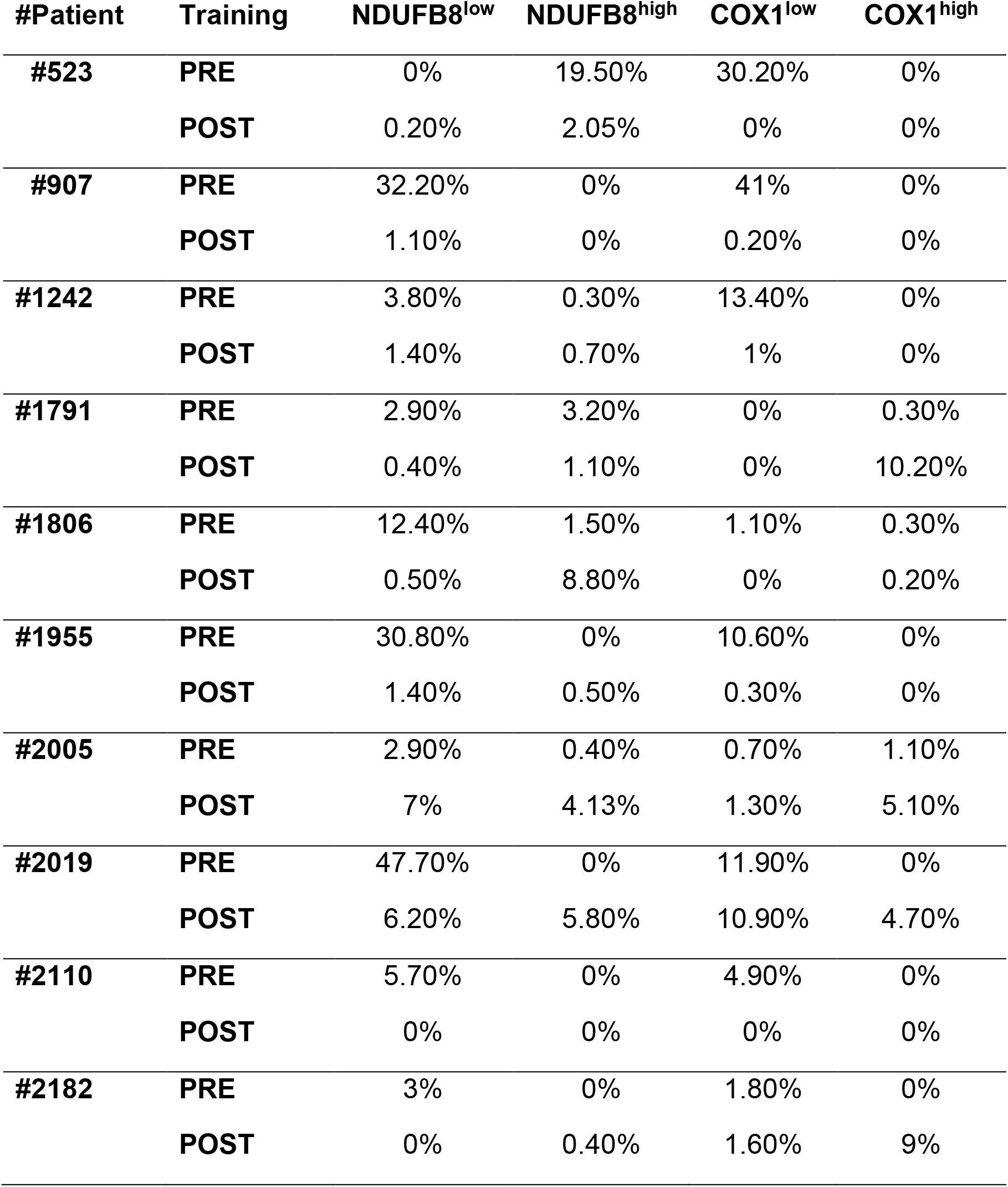
Classification of fibers across the DM1 cohort. Fibers with low and high level of NDUFB8 and COX1 are reported in percent (%) for each sample of the DM1 cohort. Abbreviations: POST, post-exercise; PRE, pre-exercise.

**Figure 4:**
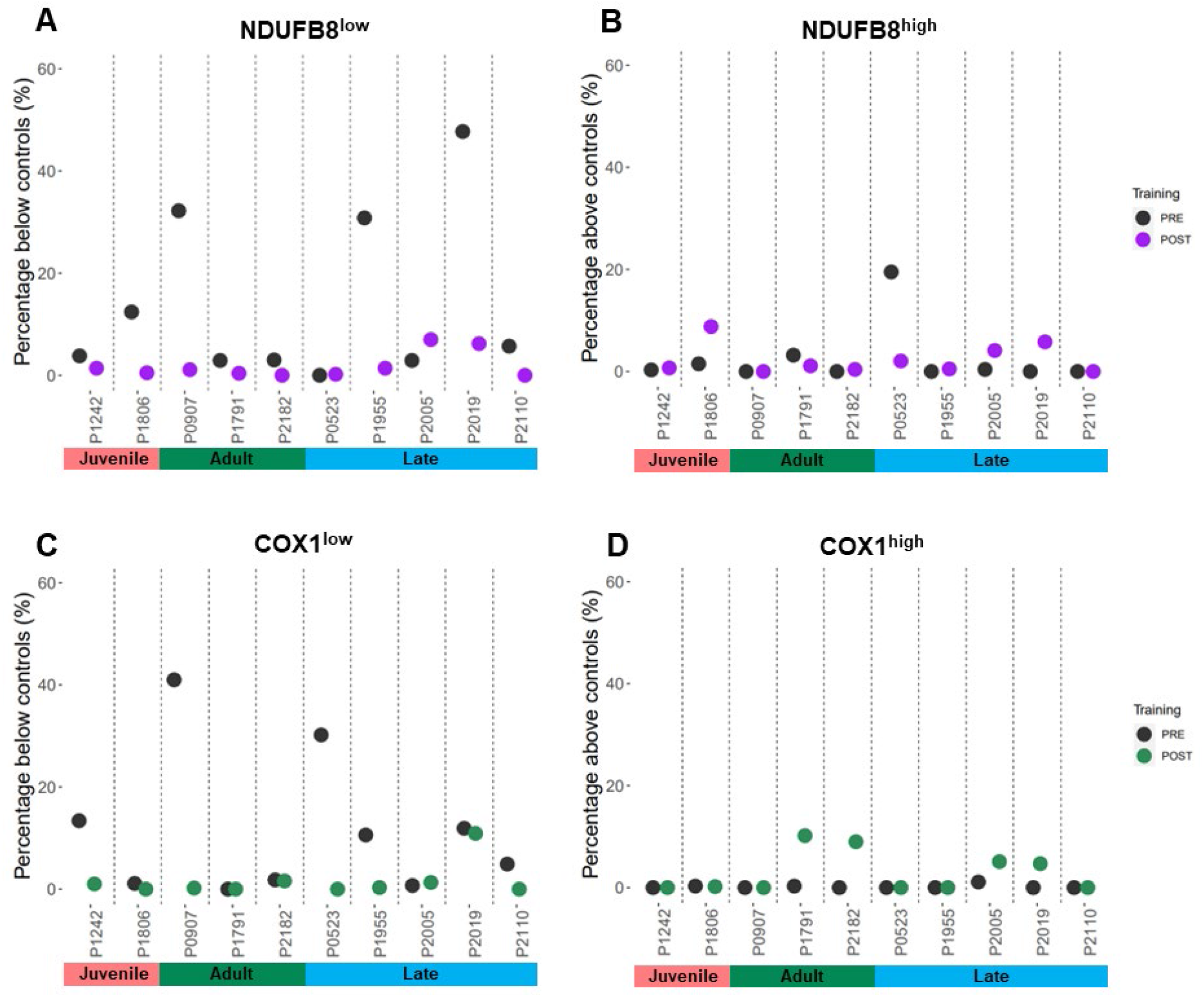
Overview of proportion of fibers with NDUFB8 and COX1 low or high in DM1 cohort. QIF data were analyzed by the 95% predictive interval model. Proportion of fibers are reported as proportion below controls with low level fibers for NDUFB8 (A) and COX1 (C), or as proportion above the controls with high level fibers for NDUFB8 (B) and COX1 (D).

### 3.4 Exercise-induced OXPHOS effects

To assess any significant change in the proportion of fibers^low^ and fibers^high^ after strength training, the changes between post- and pre-exercise cases were estimated by calculating delta (Δ). All patients, except #1242, #523 and #2005, display a significant change towards amelioration in NDUFB8^low^ fibers after exercise (p-value < 0.05, **Figure 5A**). Five patients (#1242, #907, #523, #1955, and #2110) exhibit an improvement in the proportion of COX1^low^ fibers (p-value < 0.05, **Figure 5C**). Furthermore, after exercise a significant increase in both NDUFB8^high^ (n=3) and COX1^high^ (n=4) proportion of fibers was observed (p-value < 0.05, **Figure 5B-D**). Overall, after strength training every patient in the DM1 cohort displays a significant change in at least one of the four classes of fibers identified (NDUFB8^low^, NDUFB8^high^, COX1^low^ and COX1^high^ fibers).

**Figure 5:**
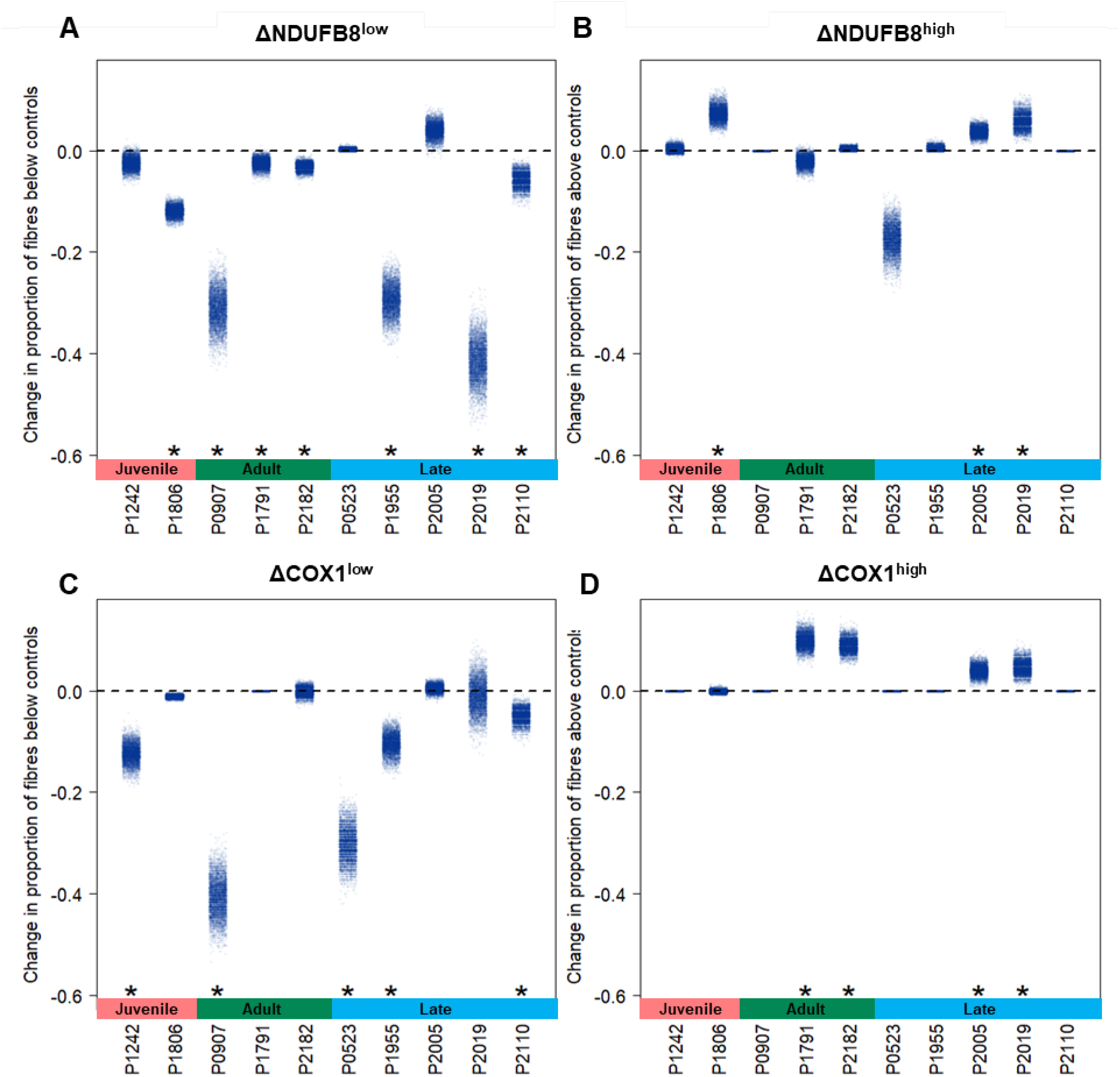
Exercise-induced effects in OXPHOS activity in DM1 cohort. Changes in the proportion of fibers (Δ) between post- and pre-exercise cases were calculated and bootstrapped to give estimates of uncertainty. Significance of changes were tested using permutation test. Changes in the proportion of fibers were analyzed in fibers below the controls with low level for NDUFB8 (A) and COX1 (C), and in fibers above with high level for NDUFB8 (B) and COX1 (D). One-sided t-test between mean values of the proportion of fibers is indicated for each patient; p-value < 0.05 (*).

### 3.5 OXPHOS deficiency as a new DM1 hallmark

To establish whether DM1 patients present specifically with isolated CI deficiency, isolated CIV deficiency or combined CI/CIV deficiency, an over-representation test was utilized to assess the proportion of COX1^low^ fibers (n=323) among NDUFB8^low^ fibers (n=528). NDUFB8^low^&COX1^low^ fibers were found to be significantly over-represented across the DM1 cohort including both pre- and post-exercise cases (p-value < 0.05, **Figure 6A**). However, a significant proportion of NDUFB8^low^&COX1^low^ fibers is over-represented in seven pre-exercise cases (#1242, #1806, #907, #2182, #1955, #2019 and #2110), and in only three post-exercise cases (#1242, #907 and #2019), where the proportion of all deficient classes of fibers decreases dramatically after exercise intervention (**Figure 6B**). Similarly, the proportion of fibers^high^ was investigated and an over-representation test was used to assess the proportion of COX1^high^ fibers (n=145) among NDUFB8^high^ fibers (n=168). Only eight NDUFB8^high^&COX1^high^ fibers were identified in patient #2005 post-exercise sample, suggesting no overall significant over-representation of this class of fibers across the DM1 cohort (**Figure 6C**).

**Figure 6:**
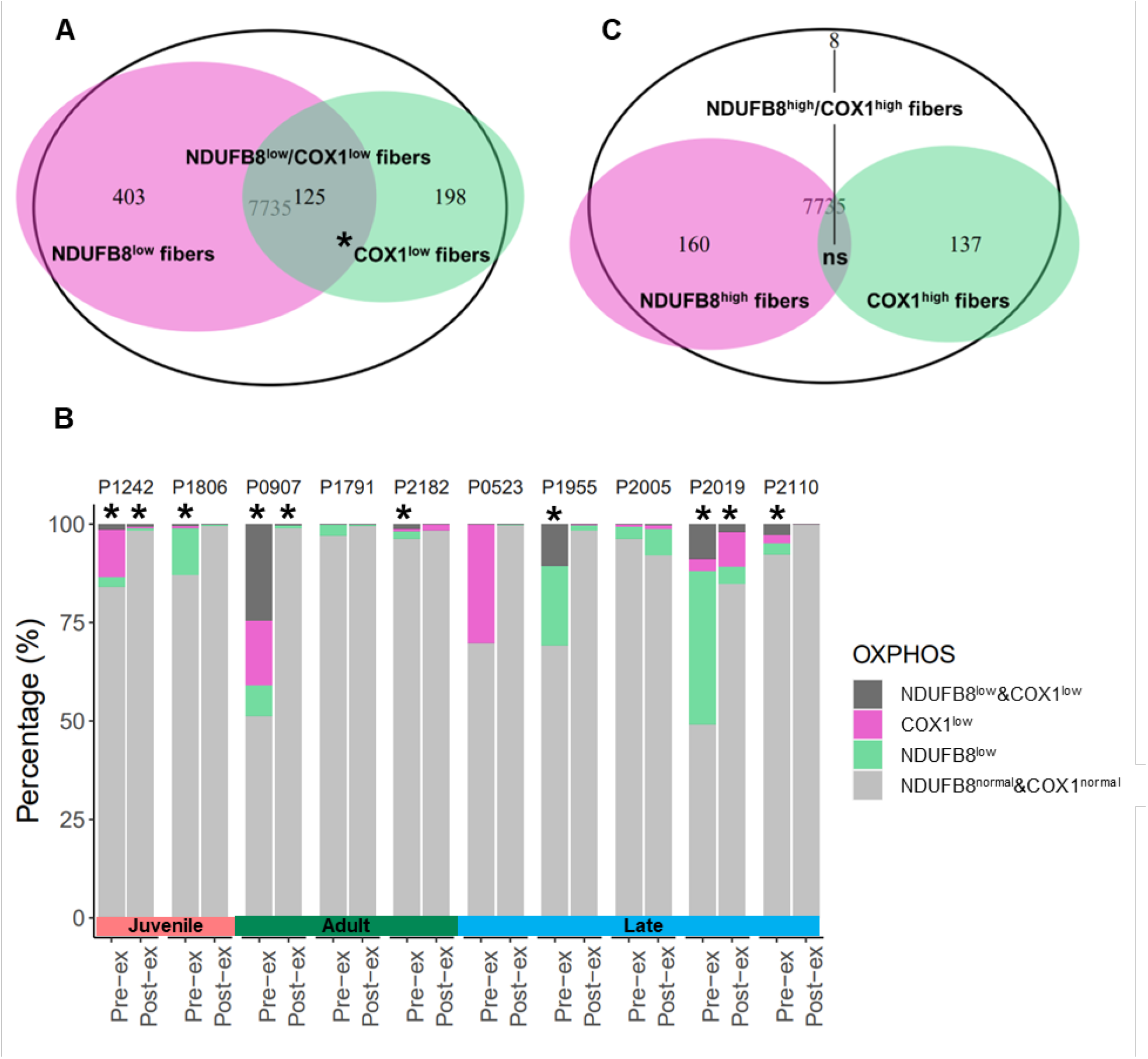
OXPHOS deficiency is a secondary trait of DM1 pathology. The total number of SKM fibers from the DM1 cohort includes 7,735 fibers, excluding the healthy control subjects. Some of the OXPHOS deficient fibers present with isolated NDUFB8 deficiency (NDUFB8^low^, n=528), isolated COX1 deficiency (COX1^low^, n=323) or both NDUFB8 and COX1 deficiency (NDUFB8l^ow^&COX1^low^, n=125) (A). Pre- and post-exercise pairs of samples are shown displaying the percent of fibers NDUFB8^low^, COX1^low^ and NDUFB8^low^&COX1^low^ (B). Some fibers present with high level of NDUFB8 (NDUFB8^high^, n=168), high level of COX1 (COX1^high^, n=145), and with high level of NDUFB8 and COX1 (NDUFB8^high^&COX1^high^, n=8) (C). Over-representation test was performed, p-value < 0.05 (*).

## 4. Discussion

The aim of this study was to investigate any potential OXPHOS dysfunction and OXPHOS protein changes after 12-week strength training in a cohort of DM1 patients [34]. Aerobic exercise training has been demonstrated to ameliorate OXPHOS deficiency in DM1 patients [37]. However, for the first time in the context of strength training, we investigated OXPHOS in SKM tissue from DM1 patients. Strikingly, we showed that DM1 SKM biopsies present with an overall decrease in mitochondrial mass when compared to healthy controls. Seven of 10 patients showed isolated or combined NDUFB8 and/or COX1 deficiency compared to healthy controls, findings which strongly resemble mitochondrial defects observed in mitochondrial myopathy patients with primary mitochondrial dysfunction [40]. Indeed, clinical presentations of DM1 and mitochondrial disease patients are variable and overlapping, usually presenting as systemic neuromuscular and metabolic disorders [41, 42]. As mentioned, DM1 pathogenicity has been associated with mitochondrial dysfunction and metabolic impairment [18, 19, 22, 26]. Of note, SKM-specific mitochondrial dysfunction has also been described in other neuromuscular disorders, such as inclusion body myositis [43, 44], dysferlin-related myopathy [45] or myofibrillar myopathy [46].

In our study, a 12-week strength training program was sufficient to induce significant changes in OXPHOS defects in SKM of DM1 patients. A significant increase in mitochondrial mass was observed in six of 10 patients independently of the phenotypical presentation (#1242 and #1806, juvenile; #1791 and #907, adult; #523 and #2110, late) (**Figure 3B**). Strikingly, these patients, excluding case #2110, present with duration of disease longer than 20 years (**Table 1**) and reported greater improvements in the ability to lift weights after strength training compared to other participants [34]. Patients #2182, #1955, #2005 and #2019 and did not exhibit an increase in mitochondrial mass, but rather a significant decrease after exercise intervention. It is important to note that VDAC1 mean value of pre-exercise cases in these patients was similar or even higher than the healthy controls mean, suggesting that the mitochondrial mass was not affected at baseline. Similarly, some of the smallest changes in lifting weights after strength training were reported in patients #1955, #2019 and #2182, who were some of the strongest at baseline [34]. These patients also show a duration of disease smaller than 20 years compared to the rest of the DM1 cohort. Altogether, these observations strongly suggest that irrespective of which phenotypical presentation patients are classified in, there is space for improvement both in terms of augmenting muscle strength [34] and increasing SKM mitochondrial mass.

A significant rescue of mitochondrial deficiency of both NDUFB8 and COX1 was observed in all DM1 patients after strength training. After strength training intervention, significant changes were observed in at least one of the four classes of fibers (NDUFB8^low^, NDUFB8^high^, COX1^low^ and COX1^high^) for each patient, indicating that the 12-week strength training program is sufficient to induce protein level changes in SKM of DM1 patients. Similarly, strength training has been demonstrated to induce a significant increase in COX1 level in healthy individuals [47, 48], and in mitochondrial myopathy patients [36]. Furthermore, an ongoing proteomics analysis performed by colleagues, demonstrated that in the DM1 cohort 12-week strength training induces substantial changes in many other mitochondrial proteins involved in the structure and function of OXPHOS complexes I and III (NDUFS2, NDUFS5, NDUFS8, CYC), beta-oxidation and Krebs cycle (IDH3A, SUCA, CISY, SUCB1, DECR), mitochondrial translation (EFTU), mitophagy and chaperonins (PHB2, CH60, CH10), antioxidation and detoxification (PRDX5, AL1B1) [49]. Mikhail and colleagues similarly showed how mitochondrial function might play a prominent role in the amelioration of physical capabilities and SKM performance in DM1 disease [37].

In DM1 patients, the CTG triplet repeat expansion is used as a disease severity marker. The rescue of NDUFB8 and COX1 however is achieved in an independent manner to CTG triplet repeat expansion, meaning that even patients with the highest CTG triplet repeat expansions (#1242 and #1806) showed improvements in NDUFB8 and COX1 deficiency after strength training intervention. Indeed, the proportion of fibers with combined NDUFB8 and COX1 deficiency (NDUFB8^low^&COX1^low^) is significantly higher in pre-exercise cases compared to post-exercise case. Moreover, #1242, #907 and #2019 post-exercise display a decrease of all deficient classes of fibers compared to the proportion observed before exercise. The changes in CI and CIV deficiency are independent of the phenotypical presentation classification, implying that overall OXPHOS defects might be a secondary trait in DM1 disease pathology, but still a DM1 hallmark that may be useful to determine the effects of exercise training intervention [34, 37]. Murine studies identified a substantial decrease of RNA foci after aerobic exercise highlighting a direct link between exercise and pathophysiological mechanisms [50]. Future investigations will underpin whether the same effects are observed in DM1 patients after 12-week strength training.

After exercise intervention, hypertrophy of both type I fibers and type II fibers was observed in subsets of DM1 patients [34], and as mentioned above, six of 10 DM1 patients displayed an increased SKM mitochondrial mass. Interestingly, changes observed in the minimal Feret’s diameter [34] and in the mitochondrial mass after exercise follow a positive, although not significant, correlation (R=0.38, p=0.28; data not shown). It is known that strength training results in hypertrophy of SKM fibers, which could be mediated by the PGC-1α4 isoform, the master regulator of mitochondrial biogenesis [51, 52]. The potential induction of PGC-1α4 isoform by strength training might explain the partial OXPHOS rescue observed. Further investigations will underpin on one side the nuclear factors that may be playing a role in the modulation of exercise responses based on inter-individual variability [53], and on the other the exercise-induced molecular targets driving the increase in SKM fiber size [34] and the rescue of OXPHOS deficiency in DM1 patients.

## Limitations

To date, this study includes the largest cohort of DM1 patients enrolled in a strength training program [33, 34]. The stringent recruitment criteria and the voluntary basis of enrolment played a crucial role in the process. All the recruited patients committed fully to the study, scoring an attendance between 90.5% and 100%, which made all the collected results extremely reliable [34]. However, comparing repeated biopsies over time necessarily involves the analysis of different myofibers, although samples were collected from the same individual and from the same tissue. This implies a certain degree of natural intra-muscle variability, which is a topic particularly involved with mitochondrial dysfunction [54]. The number of participants did not allow a meaningful statistical correlation between clinical outcomes and biological changes to be performed (data not shown). Overall, the main limitation of the study is that only male individuals were enrolled in the strength training program [34]. While this might have narrowed the inter-individual variability linked to sex, it will be important to assess the effects of strength training on a cohort of DM1 female individuals, for which an ongoing trial has been carried over (NCT05400629).

## 5. Conclusions

In conclusion, we corroborated that DM1 patients present with reduced mitochondrial mass and mitochondrial OXPHOS deficiency in both CI and CIV in SKM tissue, which suggests mitochondrial dysfunction as a DM1 hallmark. For the first time, we demonstrated that a supervised 12-week strength training program is sufficient to both increase mitochondrial mass in six of 10 patients, and partially rescue OXPHOS defects in all patients. However, the mitochondrial OXPHOS changes observed are independent from the phenotypical presentation, hence the length of the CTG triplet repeat expansion. This implies not only that mitochondrial dysfunction might be a secondary trait of DM1 pathology, but also that improvement is independent from the CTG triplet repeat expansion carried by DM1 patients. Strength training could be systematically used as an inexpensive way to treat DM1 patients with the aim of restoring physical capabilities, ameliorating SKM performance and slowing disease progression.

## Data Availability

All data produced in the present study are available upon reasonable request to the authors

https://github.com/VDLNCL/DM1-mitochondria

## 6. Author contributions

E.D. and M-P.R. were responsible for collection of the skeletal muscle samples and training of the DM1 patient cohort. V.D.L. was responsible for the study conception and design. V.D.L., C.L. and T.B.G. performed the statistical analysis of the data generated during the study. M-P.R. and E.D. ensured consistency between the previous study [34] and the study here presented. V.D.L., C.L., M-P.R., E.D. and A.E.V. were involved in the interpretation of the data. A.E.V., G.S.G., O.M.R. and H.A.L.T. were responsible for the supervision of the project as part of a Ph.D. program. V.D.L. was responsible for drafting the manuscript. All authors evaluated the manuscript, and reviewed the final publication.

## 7. Declaration of competing interest

The authors have no conflict of interest to report.

## 8. Acknowledgments including sources of support

The authors of this study would like to thank all the participants, who decided to enroll in the strength training program and to donate their samples to research. We would like to thank all the colleagues who have been involved in the recruitment of patients, organization of the strength training program and in the collection of the samples. Dre Cynthia Gagnon, who provided support with her research team, the Groupe de recherche interdisciplinaire sur les maladies neuromusculaires. Hélène Simard, who recruited all the participants in this study. Dre Catherine Savard and Dre Mylène Perron, who performed the muscle biopsies. Dr Richard Debigaré, who gave support for the interpretation of the results. We would like to thank both Université du Québec à Chicoutimi, in particular the sports center for the access to equipment and rooms for training and evaluations. We would like to thank Newcastle University, particularly the Wellcome Centre for Mitochondrial Research for the access to the lab and equipment for the analysis of the skeletal muscle biopsies, and the Bioimaging Unit for the support provided.

## 9. Funding

This project was funded by the Fondation du grand défi Pierre Lavoie and the Wellcome Centre for Mitochondrial Research. Dr Valeria Di Leo was supported by a Wellcome Ph.D. project match funded by Newcastle University (C0163N3028). Dr. Elise Duchesne is supported by a Chercheur boursier Junior 1 salary award from the Fonds de recherche du Québec-santé (FRQS-311186). Dr Amy E. Vincent is supported by a Sir Henry Wellcome Postdoctoral Fellowship (215888/Z/19/Z). Marie-Pier Roussel holds a Ph.D. study grant from the Fonds de recherche du Québec-santé (FRQS-35965).

## 10. Research data

Data linking to github repository (https://github.com/VDLNCL/DM1-mitochondria). Data statement: Roussel *et al*. 2020.

## Notes

### Competing Interest Statement

The authors have declared no competing interest.

### Clinical Trial

NCT04018820

### Funding Statement

This study was funded by the Fondation du grand defi Pierre Lavoie and the Wellcome Centre for Mitochondrial Research. Dr Valeria Di Leo was supported by a Wellcome Ph.D. project match funded by Newcastle University (C0163N3028). Dr. Elise Duchesne is supported by a Chercheur boursier Junior 1 salary award from the Fonds de recherche du Quebec-sante (FRQS-311186). Dr Amy E. Vincent is supported by a Sir Henry Wellcome Postdoctoral Fellowship (215888/Z/19/Z). Marie-Pier Roussel holds a Ph.D. study grant from the Fonds de recherche du Quebec-sante (FRQS-35965).

### Author Declarations

Ethical approval was granted by Newcastle University Research Policy Intelligence and Ethics Team, Research Strategy & Development. Here below the approval: "Based on your answers, the University Ethics Committee grants its approval for you to start working on your project. Please be aware that if you make any significant changes to your proposal then you should complete this form again, as further review may be required. This confirmation may be used within a research portfolio as evidence of ethical approval. Please note: this confirmation will be the only correspondence you should expect to receive as evidence of ethical approval. There will be no other confirmation provided. You may now proceed with research".

