## Supplemental Figure for "Strength training rescues mitochondrial dysfunction in skeletal muscle of patients with myotonic dystrophy type 1"

**Supplementary material**

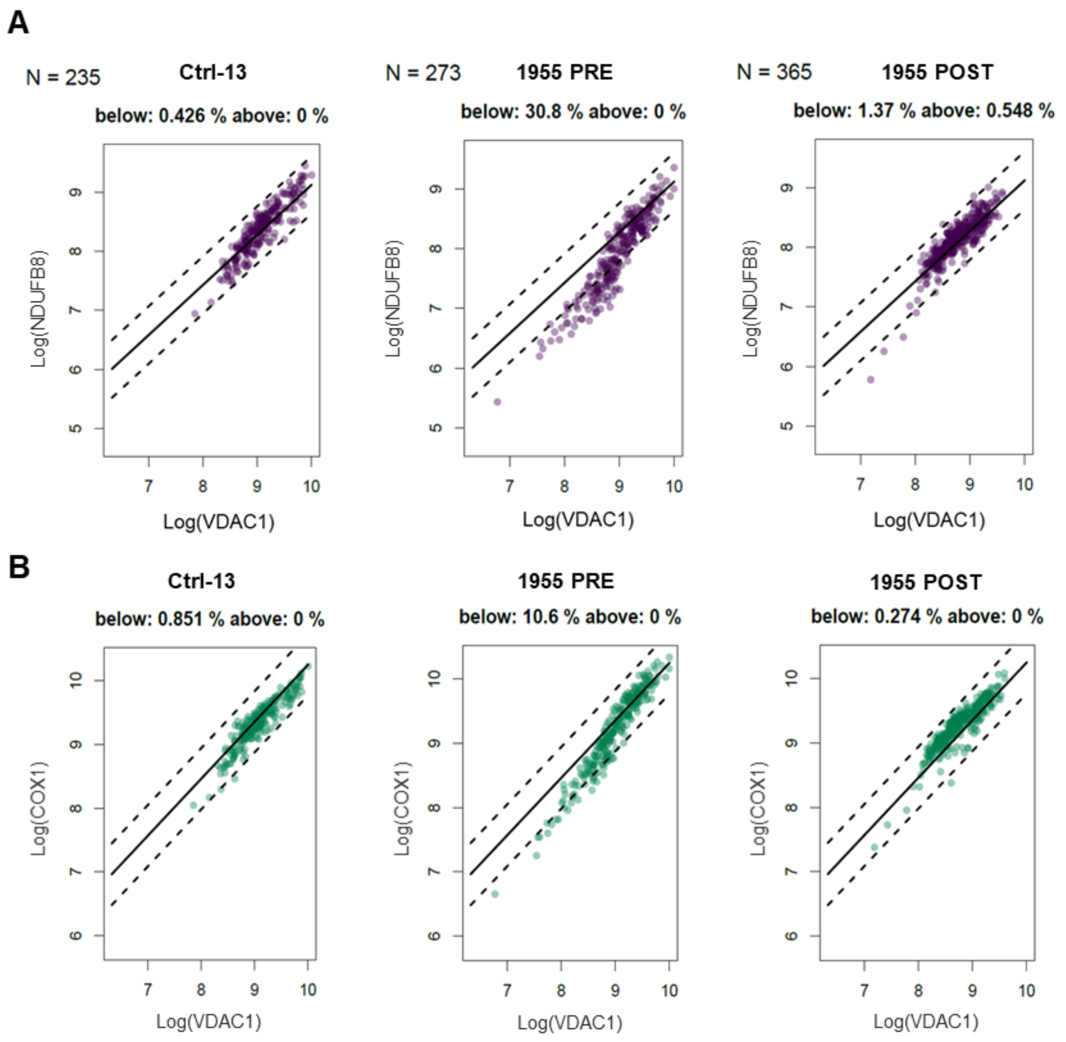


***Supplementary Figure 1: 2DMito plots report derived from QIF of DM1 cohort.***

The 2DMito plots profiles are from healthy control Ctrl-13, 1955 PRE (pre-exercise) and 1955 POST (post-exercise) cases from the DM1 cohort. The patients' fibers classification was conducted through the linear regression and 95% predictive interval model. The 2DMito plots display the linear relationship between the log transformed intensity values of VDAC1 (x axis) and NDUFB8 for complex I (A) or COX1 for complex IV (B) (y axis), respectively. Each 2DMito plot reports the total number of fibers included in the analysis, the percent of fibers below and above the predictive intervals. The fibers below the predictive intervals are classified as deficient or with low level ($fibers^{low}$), the fibers between the predictive intervals as normal fibers, whereas the fibers above the predictive intervals as overexpressing or with high level ($fibers^{high}$).
